# Cellular and humoral immunity to SARS-CoV-2 infection in multiple sclerosis patients on ocrelizumab and other disease-modifying therapies: a multi-ethnic observational study

**DOI:** 10.1101/2022.01.10.22268752

**Authors:** Ilya Kister, Yury Patskovsky, Ryan Curtin, Jinglan Pei, Katherine Perdomo, Zoe Rimler, Iryna Voloshyna, Marie I. Samanovic, Amber R. Cornelius, Yogambigai Velmurugu, Samantha Nyovanie, Joseph Kim, Ethan Tardio, Tamar E. Bacon, Lana Zhovtis Ryerson, Pranil Raut, Rosetta Pedotti, Kathleen Hawker, Catarina Raposo, Jessica Priest, Mark Cabatingan, Ryan C. Winger, Mark J. Mulligan, Michelle Krogsgaard, Gregg J. Silverman

## Abstract

**Objective:** To determine the impact of MS disease-modifying therapies (DMTs) on the development of cellular and humoral immunity to SARS-CoV-2 infection.

**Methods:** MS patients aged 18-60 were evaluated for anti-nucleocapsid and anti-Spike RBD antibody with electro-chemiluminescence immunoassay; antibody responses to Spike protein, RBD, N-terminal domain with multiepitope bead-based immunoassays (MBI); live virus immunofluorescence-based microneutralization assay; T-cell responses to SARS-CoV-2 Spike using TruCulture ELISA; and IL-2 and IFNγ ELISpot assays. Assay results were compared by DMT class. Spearman correlation and multivariate analyses were performed to examine associations between immunologic responses and infection severity.

**Results:** Between 1/6/2021 and 7/21/2021, 389 MS patients were recruited (mean age 40.3 years; 74% female; 62% non-White). Most common DMTs were ocrelizumab (OCR) - 40%; natalizumab - 17%, Sphingosine 1-phosphate receptor (S1P) modulators −12%; and 15% untreated. 177 patients (46%) had laboratory evidence of SARS-CoV-2 infection; 130 had symptomatic infection, 47 - asymptomatic. Antibody responses were markedly attenuated in OCR compared to other groups (p≤ 0001). T-cell responses (IFNγ were decreased in S1P (p=0.03), increased in natalizumab (p<0.001), and similar in other DMTs, including OCR. Cellular and humoral responses were moderately correlated in both OCR (r=0.45, p=0.0002) and non-OCR (r=0.64, p<0.0001). Immune responses did not differ by race/ethnicity. COVID-19 clinical course was mostly non-severe and similar across DMTs; 7% (9/130) were hospitalized.

**Interpretation:** DMTs had differential effects on humoral and cellular immune responses to SARS-CoV-2 infection. Immune responses did not correlate with COVID-19 clinical severity in this relatively young and non-disabled group of MS patients.

## Introduction

Multiple Sclerosis (MS) is treated with disease-modifying therapies (DMTs), some of which may impair immune responses to the pandemic SARS-CoV-2 infection. The commonly used anti-CD20 therapies (aCD20) are associated with reduced antibody titers following SARS-CoV-2 infection and vaccination^1–10^, likely due to depletion of peripheral B cells that would otherwise be available for recruitment into germinal centers for antigen-mediated activation and clonal expansion. T-cell compartment is relatively unaffected by aCD20 as only a small subset of CD20-bearing CD3+ lymphocytes are removed by aCD20.^11, 12^ Overall, T-cells counts and functional responses remain intact, ^13, 14^ and, accordingly, Spike protein-specific T-cell responses to COVID-19 vaccination in aCD20-treated patients are robust.^11, 15–19^ T-cell responses following natural infection in aCD20-treated patients have received less attention, but are an active area of investigation.^20, 21^ There is very limited data on immune responses to SARS-CoV-2 infection under other commonly used DMTs, such as sphingosine-1-phosphate receptor modulators (S1P), which interfere with T-cell egress from lymphoid tissue, and fumarates, which induce mild-moderate lymphopenia.^20, 22, 23^ Understanding the impact of DMTs on the immune response to SARS-CoV-2 infection is critical for counseling MS patients about COVID-19 risks and determining whether a patient who experienced COVID-19 on a particular DMT is likely to derive a similar degree of protective immunity as untreated individuals. ^24–27^

To address the knowledge gaps, we designed a prospective study with the goals of: 1. determining the impact of ocrelizumab (OCR) and other DMTs on the development of cellular and humoral immune memory to SARS-CoV-2 natural infection; 2. characterizing the relationship between humoral and cellular post-infection immune responses in patients with and without peripheral B-cell depletion; 3. investigating the relationship between the clinical severity of COVID-19 and immune responses to SARS-CoV-2 in MS patients with different DMTs. We recruited a large, ethnically-diverse group of MS patients from the NYU MS Care Center in New York City, New York, one of the epicenters of the COVID-19 pandemic in 2020-21,^28, 29^ and comprehensively characterized humoral and cellular responses to SARS-CoV-2 using several complementary antibody and T-cell SARS-CoV-2 - specific assays. A notable strength of our study is the inclusion of a large number of non-White patients - over 60% of all patients, which allowed us to investigate immune responses to SARS-CoV-2 in MS patients from underrepresented racial/ethnic groups.

## Methods

### Study population

Patients seen for routine visits at the NYU Multiple Sclerosis Comprehensive Care Center in New York City (New York) were invited to participate if they had clinician-diagnosed MS (revised 2017 McDonald criteria);^30^ were treated with an FDA-approved DMT for MS, or were on no treatment; were aged 18 to 60; had Expanded Disability Status Scale (EDSS) score of 0 (normal) to 7 (wheelchair-bound). Exclusion criteria were: concurrent immunosuppressive therapy; active systemic cancer; primary or acquired immunodeficiency (unrelated to DMT); active drug or alcohol abuse; aCD20 therapy other than OCR; uncontrolled diabetes mellitus; end-organ failure (cardiac, pulmonary, renal, hepatic); systemic lupus erythematosus or other systemic autoimmune disease. Patients were also excluded if they received high-dose oral or parenteral corticosteroids, intravenous immunoglobulin (IVIG), plasmapheresis (PLEX), or convalescent plasma or polyclonal antibody treatments for COVID-19 within three months of sample collection; or if they had COVID-19 symptom onset or tested positive by SARS-CoV-2 real-time PCR within two weeks of sample collection. At the time of sample collection all patients were unvaccinated for COVID-19.

All patients were interviewed by a trained research coordinator with a structured instrument. Patients were queried about each of COVID-19 symptoms listed in CDC clinical case definition^31^ and any COVID-19 exposures from February 2020 to the time of enrollment; commercial SARS-CoV-2 test results (PCR or Antibody) prior to enrollment; COVID-19 treatments and vaccinations; MS treatment at the time of enrollment and infection (where applicable). Electronic medical records were reviewed for COVID-19 and MS-relevant information. COVID-19 history *at enrollment* was classified as ‘Laboratory-supported COVID infection at enrollment’ if the patient met CDC clinical definition for COVID-19 *and* had positive commercial SARS-CoV-2 PCR or antibody test at any time prior to enrollment. If the patient met the CDC clinical case definition for COVID-19 but did not have commercial laboratory confirmation at the time of enrollment, their status was designated as ‘Suspected COVID-19 infection on enrollment.’ Patients without clinical symptoms to suggest prior COVID-19 were classified as ‘Non-suspected’ for COVID-19 infection. Patients’ *final* SARS CoV-2 infection status (previously infected *vs.* non-infected) was determined based on laboratory evidence of infection prior to enrollment or serologic tests conducted during the study as described in the next section.

### Serological analyses

#### Patients’ serologic status was assessed using three different methods

1. Electro-chemiluminescence immunoassay using the Elecsys® platform (Roche Diagnostics), measuring antibodies to Nucleocapsid (N) (qualitative) and Receptor Binding Domain (RBD) of Spike (S) protein (quantitative). All samples were processed and measured by a specialized laboratory according to the manufacturers’ instructions. Values ≥1.0 U/mL were interpreted as ‘positive’ for anti-N SARS-CoV-2 antibodies. For anti-Spike Abs, values of > 0.4 U/mL were considered ‘positive’, and those below the lower limit of quantification of the assay (<0.4 U/mL) were considered ‘negative’ and set to 0.4 U/mL, as per the manufacturer’s recommendations ^32^. The levels of antibodies were expressed in U/mL, which are considered equivalent to Binding Antibody Units (BAU)/mL (Elecsys S Units = 0.972 x BAU), as defined by the first World Health Organization (WHO) International Standard for anti-SARS-CoV-2 immunoglobulin (NIBSC code 20/136).^32^

2. NYU proprietary custom Multiepitope Bead-based Immunoassay (MBI), which measures antibody responses to three recombinant proteins (Wuhan variant total Spike, RBD and the S amino-terminal domain (NTD); Sino Biological cat no. 40590-V08B, 40592-V08B, 40591-V49H-B, respectively), using control analytes of Human serum albumin (HSA), tetanus toxoid and anti-human IgG (Jackson Immunoresearch Inc.) coupled to commercial paramagnetic beads (MagPix, Luminex), as adapted from the manufacturer’s instructions as previously described.^33, 34^ Positivity of individual MBI was set as the 3SD above the mean of pre-pandemic healthy adult controls. Assay reactivity was also confirmed with non-autoimmune serum from individuals with PCR-documented prior COVID-19 infection (samples provided by the NYU COVID-19 Bio Repository). MBI data for S, RBD, and NTD for healthy control and COVID-19 patient specimens and respective positivity cut-offs are shown in the Supplemental Figure 1. Serologic confirmation of prior SARS-CoV-2 infection (’MBI seropositive’) was defined conservatively as *two or more independent* S, RBD, and NTD positive assay results.

3. SARS-CoV-2 viral neutralization activity of plasma was measured in an immunofluorescence-based assay that detects the neutralization of infectious virus (SARS-CoV-2 isolate USA-WA1/2020 (NR-52281, GenBank accession no. MT233526) in cultured Vero E6 cells (African Green Monkey Kidney; ATCC #CRL-1586) as described in detail in ^35^. All SARS-CoV-2 infection assays were performed in the BSL3 facility of NYU Grossman School of Medicine (New York, NY).

### Assays of SARS-CoV-2-specific T-cell response

T-cell responses to SARS-CoV-2 Spike protein were assessed in whole blood samples with TruCulture® stimulation system (Myriad RBM) according to the manufacturer’s instructions. Whole blood samples were incubated for 48 hours at 37° C. Collected supernatants were analyzed with IFNγ and IL-2 ELISA assay for measuring cytokine production following manufacturer’s protocol (Fisher Scientific, Cat # ENEHIFNG and 50-112-5363, respectively). The response for the TruCulture system was conservatively defined as positive if *both* IFNγ and IL-2 assays were at the level of ≥1 pg/ml.

Cellular responses to SARS-CoV-2 were also characterized by quantification of IFNγ and IL-2 producing cells by ELISpot for a subset of patients to corroborate results obtained with the TruCulture system. Briefly, peripheral blood mononuclear cells (PBMCs) were in vitro stimulated with 1 µg/peptide/ml SARS-CoV-2 peptide pool, consisting of Spike, N, and M proteins15-mer peptide *PepTivator* libraries (*Miltenyi Biotec*) for 48 hours. The number of activated T-cells were detected with *ImmunoSpot* IFNγ and IL-2 kits (Cellular Technology Limited, Cat # hIFNgp-2M/10 and hIL2p-2M/10) according to the manufacturer’s instructions, and spots were counted using a CTL S6 EM2 ELISpot reader (Immunospot). Positive results were confirmed by repeated ELISpot assays. The results were expressed as spot-forming units (s.f.u.) per 10^6^ PBMCs. Responses were considered positive if the results were at least three times the mean of the negative control wells and >25_s.f.u. per 106 PBMCs. Human CEF (CMV, Epstein-Barr virus, Influenza virus) peptide pool (3615–1) (MabTech) and 1_μg/mL phytohemagglutinin-L (PHA-L) (Sigma Aldrich) were used as positive controls. The negative control contained PBMC, and the corresponding cell culture medium was used to determine the background signal.

### Statistical analyses

All patients were included in the analyses. Descriptive summaries of the results from the immunoassays were reported for continuous and categorical variables. Results that have heavily skewed distributions were normalized by log transformation. For continuous variables, mean, standard deviation (SD), median, and range were reported. For categorical variables, counts and percentage of patients with positive results were summarized. Correlation analyses were performed using the Spearman correlation. Comparisons of endpoints were performed between patients on the various DMTs and untreated patients (no DMT). Multivariate analyses were performed to account for possible confounding characteristics, including the patients’ COVID-19 clinical severity and MS treatments. Missing data were not imputed.

The study was approved by the NYU Grossman School of Medicine IRB.

## Results

### 1. Demographic and clinical characteristics of the patients

From January 6, 2021, to July 21, 2021, 389 non-vaccinated MS patients were recruited. Demographic and clinical characteristics, DMT use, COVID-19-relevant comorbidities of the patients are shown in Table 1. The patients were relatively young (mean age: 40.3±10.8 years, range 18–60), non-disabled (68% fully ambulatory), and otherwise healthy (68% without any COVID19-relevant comorbidities). Sex ratio (74% female) and race/ethnic composition (62% non-White) of the patients are representative of our clinic population.^36^ ‘COVID history at enrollment’ was classified as ‘laboratory-supported COVID at enrollment’ in 101 patients (26% of all patients), ‘suspected COVID at enrollment’ (symptoms only) in 76 patients (20%), and ‘COVID non-suspected’ in 212 patients (54%). These three subgroups had similar demographic and clinical MS characteristics (data not shown).

**Table 1.** Demographic and clinical characteristics of patients with MS. Legend: S1P receptor modulators included Fingolimod (Gilenya), Siponimod (Mayzent), Ozanimod (Zeposia); Fumarates include Dimethyl Fumarate (Tecfidera), diroximel fumarate (Vumerity); Interferon β included Interferon beta-1a (Avonex, Rebif) and Interferon beta-1b (Betaseron). *COVID-relevant comorbidities included: hypertension, chronic obstructive pulmonary disease, cardiovascular disease, diabetes mellitus, obesity, sickle cell disease, chronic kidney disease, chronic liver disease and (non-skin) cancer. Abbreviations: DMT, disease-modifying therapy; MS, multiple sclerosis; S1P, sphingosine 1-phosphate receptor modulators; SD –standard deviation Q1 (quartile 1) and Q3 (quartile 3) represent median of the lower and upper half of the data.

|  | **All Patients**<br>**N=389** |
| --- | --- |
| **Age, years** |  |
| Mean (SD) | 40.3 (10.8) |
| Median (Q1, Q3) | 40.0 (32.0, 49.0) |
| **Female, n (%)** | 286 (73.5) |
| **Race/ethnicity, n (%)** |  |
| White | 147 (37.8) |
| African American/ Black | 111 (28.5) |
| Hispanic | 106 (27.2) |
| Other | 25 (6.4) |
| **DMT at enrollment, n (%)** |  |
| Ocrelizumab | 154 (39.6) |
| Natalizumab | 65 (16.7) |
| No DMT | 58 (14.9) |
| S1P | 48 (12.3) |
| Fumarates | 34 (8.7) |
| Glatiramer acetate | 11 (2.8) |
| Teriflunomide | 10 (2.6) |
| Interferon $\beta$ | 9 (2.3) |
| **Ambulatory status, n (%)** |  |
| Fully ambulatory | 265 (68.1) |
| Impaired but no assistance | 49 (12.6) |
| Assistance with cane | 45 (11.6) |
| Assistance with walker | 26 (6.7) |
| Non-ambulatory/wheelchair | 4 (1.0) |
| **Number of comorbidities\*, n (%)** |  |
| 0 | 264 (67.9) |
| 1 | 94 (24.2) |
| 2 | 24 (6.2) |
| 3 | 7 (1.8) |

### 2. SARS-CoV-2 Antibody Testing

All patients underwent serologic testing by Elecsys assay for antibodies to N and the S RBD, and by MBI for whole S protein and the S RBD and NTD components of Spike. There was a strong correlation between the individual assays for the whole Spike and Spike components by MBI (r= 0.77-0.82, p<0.0001), and between anti-RBD antibody levels by MBI and by Elecsys (r=0.69, p<0.0001). SARS-CoV-2 antibody responses did not differ by race/ethnicity (White v. Black v. Hispanics v. Other) with either Elecsys or MBI assay systems (Supplemental Table 1).

The sensitivity and specificity of two antibody assays are summarized in Table 2. The patients with ‘laboratory-supported COVID at enrollment’ are shown in the first column. Despite the more stringent definition of seropositivity for MBI (≥ of 3 independent antibody assays with >3SD above pre-pandemic means) than Elecsys (either one of the two antibodies positive), MBI had greater sensitivity for antibody detection in patients with laboratory-confirmed COVID-19 on enrollment: 92% for MBI v. 81% for Elecsys. Eight patients were considered false-negative by MBI, as they had a lab-confirmed infection before enrollment by commercial tests but were negative by MBI; all eight patients were on OCR at the time of infection.

**Table 2.** Seropositivity rates by Elecsys and MBI stratified by COVID history on enrollment. Legend: ‘Lab-supported COVID on enrollment’ is defined as ‘clinical symptoms consistent with COVID (CDC clinical case definition) and laboratory confirmation of SARS CoV-2 prior to enrollment’. ‘Suspected COVID on enrollment’ is defined as ‘clinical symptoms consistent with COVID (CDC clinical case definition) but no laboratory confirmation of SARS CoV-2 prior to enrollment’. ‘Not suspected on enrollment’ is defined as not meeting CDC clinical case definition. ‘Elecsys seropositivity’ is defined as ‘either N (nucleocapsid) or S (spike) antibody positive’ as defined by manufacturer. ‘MBI seropositivity’ is defined more stringently as ‘≥2/3 independent antibody assays >3SD over the mean pre-pandemic levels’. Abbreviations: MBI, multiepitope bead-based immunoassay; N, nucleocapsid; NTD, N-terminal domain; RBD, receptor-binding domain; S, spike.

|  | Lab-supported COVID on enrollment, n=101 | Suspected COVID on enrollment, n=76 | Not suspected on enrollment, n=212 |
| --- | --- | --- | --- |
| **Elecsys, n (%)** |  |  |  |
| N | 67<sup>a</sup> (67.0) | 12 (15.8) | 19 (9.0) |
| S | 80 (79.2) | 16 (21.1) | 23 (10.8) |
| **Seropositivity** | **82 (81.2)** | **16 (21.1)** | **23 (10.8)** |
| **MBI, n (%)** |  |  |  |
| Spike | 93 (92.1) | 32 (42.1) | 47 (22.2) |
| RBD | 95 (94.1) | 29 (38.2) | 48 (22.6) |
| NTD | 91 (90.1) | 31 (40.8) | 80<sup>b</sup> (38.5) |
| **Seropositivity** | **93 (92.1)** | **29 (38.2)** | **47 (22.2)** |

<sup>a</sup> Denominator = 100.
<sup>b</sup> Denominator = 208.

The second column of Table 2 presents the prevalence of SARS-CoV-2-specific antibodies in patients with ‘suspected COVID on enrollment’ (symptoms only) and the third column – in non-suspected cases. In the ‘suspected COVID at enrollment’ group, MBI identified 29 out of 76 as ‘MBI-seropositive’ (≥2 of 3 independent antibody assays with >3SD above pre-pandemic mean levels) and in the non-suspected group, MBI identified 47 out of 212 as MBI-seropositive. Thus, there was a total of 76 patients from ‘suspected’ and ‘non-suspected’ subsets who had strong serologic evidence of SARS-CoV-2 infection. Of these patients, half (n=38) were also positive by Elecsys. In contrast, Elecsys identified only a single case that was negative by MBI. Overall, MBI’s greater sensitivity for antibody detection proved useful in the study of immune responses to chronologically remote SARS-CoV-2 infections, especially in immunosuppressed individuals.

### 3. Defining the subset of patients with prior SARS-CoV-2 infection

In addition to the 101 patients with lab-supported COVID-19 on enrollment, we identified 76 patients who did not have laboratory testing for SARS CoV-2 prior to enrollment, but tested positive on at least 2 out of 3 independent antibody levels on MBI and were thus considered to have had SARS-CoV-2 infection. Thus, the total number of patients with test-supported SARS-CoV-2 infection in our group was 177, or 46% of all patients.

The relationship between COVID-19 status at enrollment and the final SARS-CoV-2 infection status is shown in the Sankey diagram in Figure 1. Patients with prior SARS-CoV-2 infection included 130 symptomatic cases and 47 asymptomatic cases. The asymptomatic cases constituted 27% of all SARS-CoV-2 infected patients and had similar demographic and MS-related characteristics as the symptomatic cases (data not shown). The majority of patients with asymptomatic prior infection, in addition to testing positive on 2 or more serologies by MBI, had additional collateral evidence of past infection: 23/47 (49%) tested seropositive on one or both Elecsys antibody assays; 8/47 (17%) had positive COVID-specific T-cell responses (see Section 6); and 10/47 (21%) reported a history of close exposure to COVID-19 infected individuals at home or work, or were essential personnel with a high risk of exposure.

**Figure 1.**
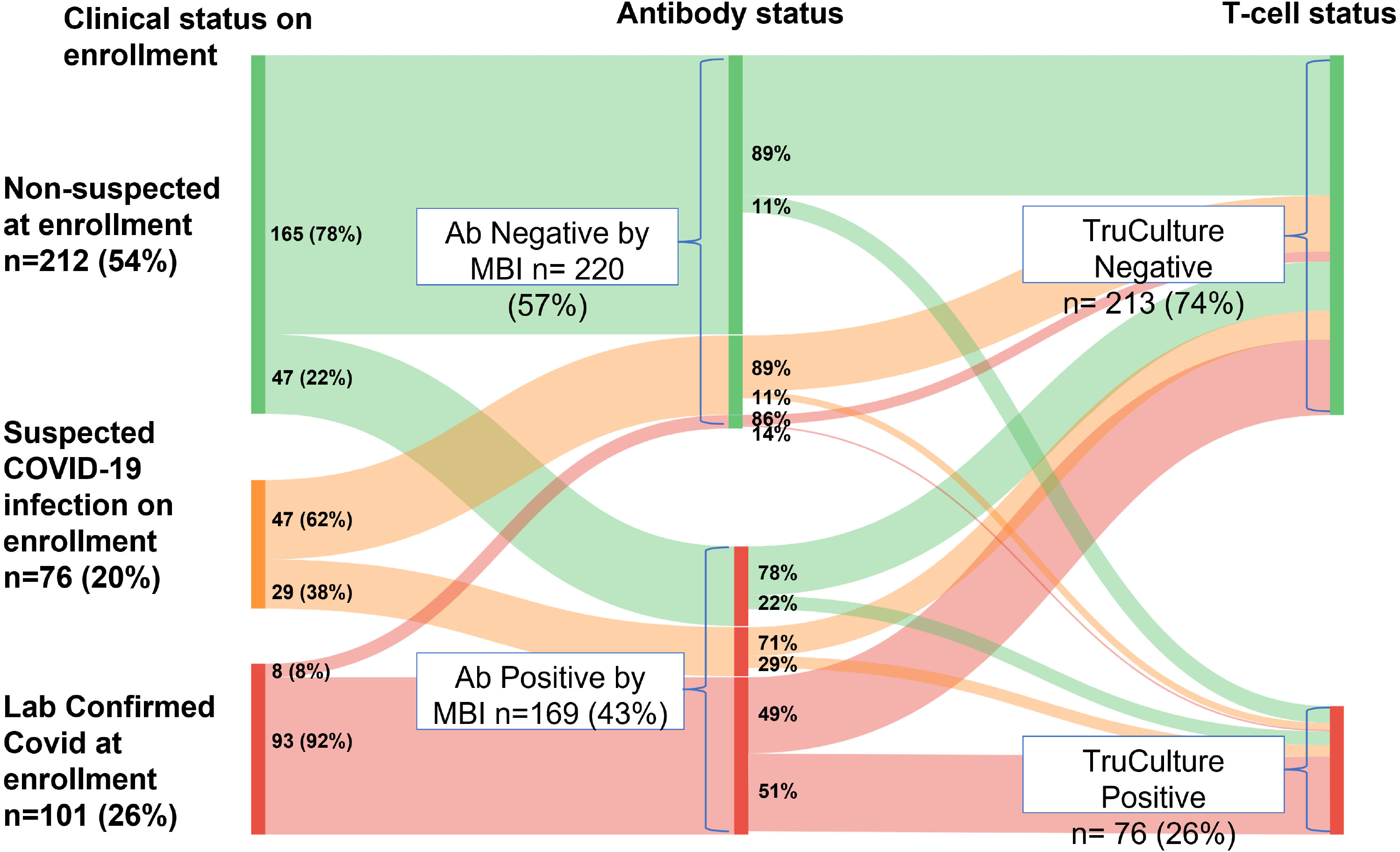
COVID-19 history at enrollment (left panel) stratified by MBI serostatus (middle) and TruCulture (right) Legend: Sankey diagram illustrates proportions of patients with ‘laboratory-supported COVID at enrollment’, ‘suspected COVID at enrollment’ and ‘COVID not suspected at enrollment’ who tested positive by MBI (middle panel) and proportion of MBI-positive and MBI-negative patients who tested positive on Truculture (see Section 6). ‘Lab-supported COVID on enrollment’ is defined as ‘clinical symptoms consistent with COVID-19 (CDC clinical case definition) and laboratory confirmation of SARS-CoV-2 prior to enrollment’. ‘Suspected COVID on enrollment’ is defined as ‘clinical symptoms consistent with COVID-19 (CDC clinical case definition) but no laboratory confirmation of SARS-CoV-2 before enrollment’. ‘Not suspected on enrollment’ is defined as not meeting CDC clinical case definition. ‘MBI seropositive’ is defined as ‘≥2/3 independent antibody assays >3SD over the mean pre-pandemic levels. ‘TruCulture positive’ was defined as *both* IFNγ and IL-2 assays were at the level of ≥1 pg/ml.

### 4. Timing of infection and clinical outcomes in patients with prior SARS-CoV-2 infection stratified by DMT

Mean time from symptomatic infection to blood collection was 34.5 ±19.0 weeks (range: 4.3–70.4 weeks). The calendar time distribution of symptomatic infections in shown in Supplemental Figure 2. The bimodal distribution matches the epidemiology of the spread of COVID-19 in NYC.^37^ All infections occurred before the spread of Delta and Omicron variants. Among the 130 symptomatic COVID-19 patients, 54 (42%) had respiratory symptoms (shortness of breath or difficulty breathing), 9 (7%) required hospitalization, and 2 were subsequently admitted to the Intensive Care Unit. The hospitalized patients (n=9) were, on average 51.2 years old and 7/9 (78%) had COVID-19-relevant comorbidities, while non-hospitalized patients were on average 39.3 years old and only 31% had comorbidities. The two patients admitted to ICU, both in their 40s, were a man with no comorbidities on dimethyl fumarate at the time of infection and a woman with three comorbidities (obesity, cardiovascular disease, and prior cancer) on S1P at the time of infection.

Demographic, clinical, and COVID-19 characteristics - number of symptoms, presence of respiratory symptoms, symptom duration, hospitalization rates - stratified by DMT class are shown in Table 3. The percentage of asymptomatic patients and COVID-19 clinical characteristics were comparable across most DMTs. However, time from infection to sample collection was much shorter for OCR (26.7 ±18.4 weeks) than all other patients (39.2 ±17.9 weeks).

**Table 3.** Clinical characteristics of MS patients with lab-confirmed COVID by DMT^a^. Legend: Lab-confirmed COVID is defined as Lab-supported COVID on enrollment and any MBI seropositive for SARS CoV2 (independent of COVID history at enrollment). S1P receptor modulators included Fingolimod (Gilenya), Siponimod (Mayzent), Ozanimod (Zeposia); Fumarates include Dimethyl Fumarate (Tecfidera), diroximel fumarate (Vumerity); Interferon β included Interferon beta-1a (Avonex, Rebif) and Interferon beta-1b (Betaseron).

|  | **OCR**<br>**n=70** | **GA**<br>**n=6** | **Interferon $\beta$**<br>**n=4** | **Fumarates**<br>**n=17** | **S1P**<br>**n=19** | **NAT**<br>**n=22** | **No DMT**<br>**n=39** | **All Patients**<br>**n=177** |
| --- | --- | --- | --- | --- | --- | --- | --- | --- |
| **COVID symptoms, n (%)** |  |  |  |  |  |  |  |  |
| Symptomatic | 49 (70.0) | 5 (83.3) | 2 (50.0) | 17 (100.0) | 12 (63.2) | 17 (77.3) | 28 (71.8) | 130 (73.4) |
| Asymptomatic | 21 (30.0) | 1 (16.7) | 2 (50.0) | 0 | 7 (36.8) | 5 (22.7) | 11 (28.2) | 47 (26.6) |
| **Age, years** |  |  |  |  |  |  |  |  |
| Mean (SD) | 37.9 (9.7) | 40.0 (13.0) | 49.0 (7.4) | 42.2 (10.7) | 43.0 (9.2) | 36.8 (11.4) | 41.9 (12.6) | 39.9 (10.9) |
| **Female, n (%)** | 53 (75.7) | 4 (66.7) | 2 (50.0) | 10 (58.8) | 16 (84.2) | 15 (68.2) | 25 (64.1) | 125 (70.6) |
| **Race, n (%)** |  |  |  |  |  |  |  |  |
| White | 27 (38.6) | 3 (50.0) | 1 (25.0) | 6 (35.3) | 6 (31.6) | 10 (45.5) | 16 (41.0) | 69 (39.0) |
| African American/Black | 14 (20.0) | 0 | 3 (75.0) | 6 (35.3) | 7 (36.8) | 6 (27.3) | 13 (33.3) | 49 (27.7) |
| Hispanic | 23 (32.9) | 2 (33.3) | 0 | 5 (29.4) | 5 (26.3) | 5 (22.7) | 9 (23.1) | 49 (27.7) |
| Other | 6 (8.6) | 1 (16.7) | 0 | 0 | 1 (5.3) | 1 (4.5) | 1 (2.6) | 10 (5.6) |
| **Number of comorbidities<sup>b</sup>, n (%)** |  |  |  |  |  |  |  |  |
| 0 | 53 (75.7) | 5 (83.3) | 2 (50.0) | 8 (47.1) | 13 (68.4) | 15 (68.2) | 22 (56.4) | 118 (66.7) |
| 1 | 15 (21.4) | 1 (16.7) | 2 (50.0) | 6 (35.3) | 5 (26.3) | 4 (18.2) | 13 (33.3) | 46 (26.0) |
| 2 | 2 (2.9) | 0 | 0 | 2 (11.8) | 0 | 3 (13.6) | 3 (7.7) | 10 (5.6) |
| 3 | 0 | 0 | 0 | 1 (5.9) | 1 (5.3) | 0 | 1 (2.6) | 3 (1.7) |
| **Symptom count, n (%)** |  |  |  |  |  |  |  |  |
| 0 (asymptomatic) | 19 (27.1) | 1 (16.7) | 2 (50.0) | 0 | 7 (36.8) | 5 (22.7) | 9 (23.1) | 43 (24.3) |
| 1-3 | 17 (24.3) | 3 (50.0) | 0 | 3 (17.6) | 4 (21.1) | 5 (22.7) | 9 (23.1) | 41 (23.2) |
| >3 | 34 (48.6) | 2 (33.3) | 2 (50.0) | 14 (82.4) | 8 (42.1) | 12 (54.5) | 21 (53.8) | 93 (52.5) |
| **Symptom duration, weeks** |  |  |  |  |  |  |  |  |
| Median (Q1, Q3) | 1.9<br>(1.0, 3.0) | 0.7<br>(0.6, 1.0) | 1.6<br>(1.0, 2.3) | 2.0<br>(1.1, 2.9) | 1.4<br>(1.0, 2.0) | 1.4<br>(1.0, 2.0) | 2.6<br>(1.0, 6.0) | 2.0<br>(1.0, 3.0) |
| **Respiratory symptoms, n (%)** | 16 (22.9) | 1 (16.7) | 0 | 12 (70.6) | 4 (21.1) | 6 (27.3) | 15 (38.5) | 54 (30.5) |
| **Hospitalization, n (%)** | 2 (2.9) | 1 (16.7) | 0 | 2 (11.8) | 1 (5.3) | 1 (4.5) | 2 (5.1) | 9 (5.1) |

<sup>a</sup> For symptomatic COVID patients, we use DMT at the time of symptoms; for asymptomatic patients – DMT at the time of enrollment. Comorbidities included: hypertension, chronic obstructive pulmonary disease, cardiovascular disease, diabetes mellitus, obesity, sickle cell disease, chronic kidney disease, chronic liver disease, and cancer (non-skin cancers only).
<sup>b</sup> COVID-relevant comorbidities: hypertension, chronic obstructive pulmonary disease, cardiovascular disease, diabetes mellitus, obesity, sickle cell disease, chronic kidney disease, chronic liver disease and (non-skin) cancer.
Abbreviations: DMT, disease-modifying therapy; GA, glatiramer acetate; NAT, natalizumab; OCR, ocrelizumab; S1P, sphingosine 1-phosphate receptor modulators.

### 5. Humoral responses among patients with prior SARS-CoV-2 infections stratified by DMT

Among patients with prior SARS-CoV-2 infection, the seropositivity rate by MBI was 100% for all DMTs except for OCR, for which the seropositivity rate was 89%. Seropositivity rates by Elecsys for non-OCR DMTs and the no-treatment group ranged from 83% to 100%, while for OCR it was only 36%.

Levels of anti-Spike antibodies by Elecsys (Figure 2A) and MBI (Figure 2B) were approximately 10-fold lower in OCR patients with prior SARS-CoV-2 compared to untreated patients with prior infection. S1P patients had significantly lower antibody levels than the untreated patients as measured by Elecsys, but the difference was not statistically significant with the MBI assay. For OCR-treated patients with a history of COVID-19, there was a non-significant trend for increased anti-SARS-CoV-2 Ab titers on MBI assay with longer time from the last OCR infusion prior to infection and infection onset (r=0.285, p=0.064).

**Figure 2:**
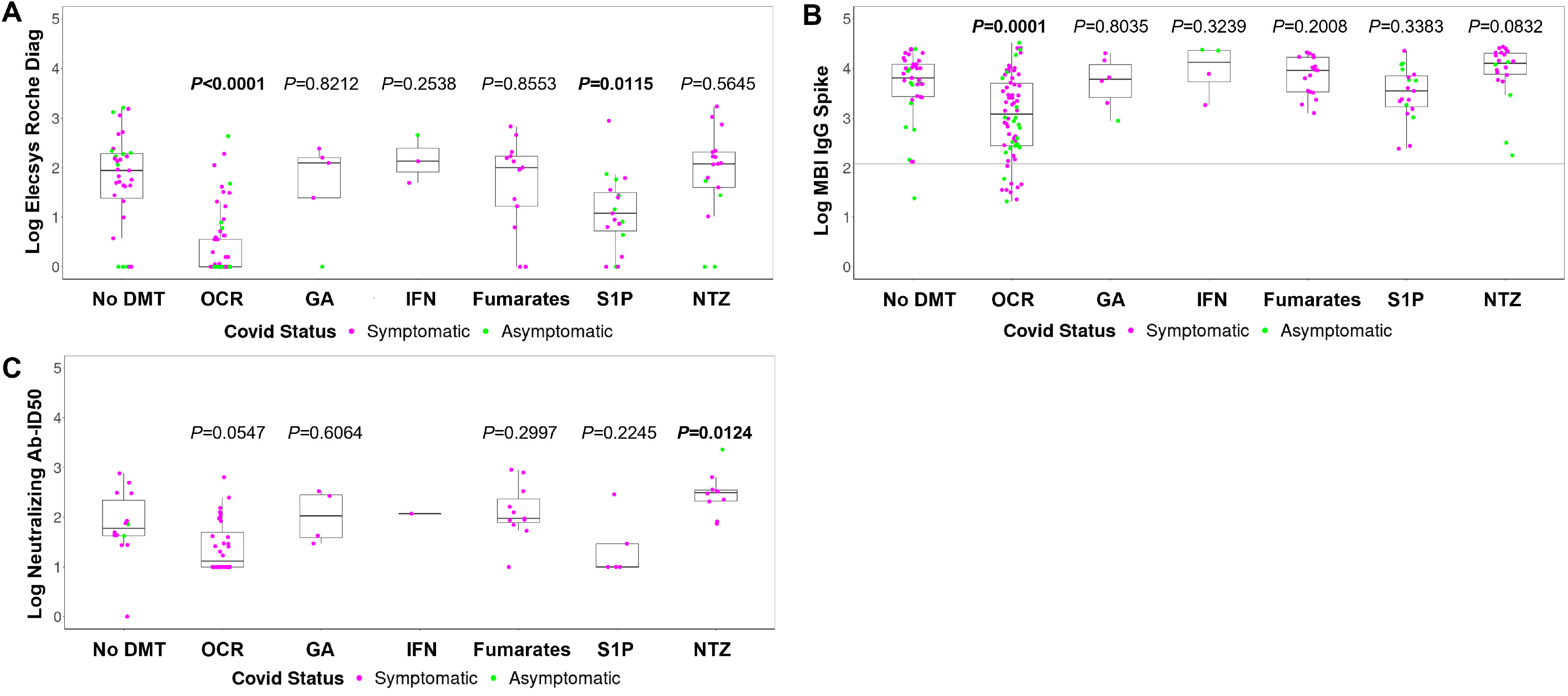
Elecsys (A), MBI (B) and Nabs levels (C) anti-Spike antibodies by DMT. Legend: Symptomatic cases are shown in magenta and asymptomatic cases – in green. Neutralizing antibody titers are shown as log10 of half-maximal inhibitory dilution (ID50). P-values <0.05 are shown in bold. Abbreviations: Ab, antibody; DMT, disease-modifying therapy; GA, glatiramer acetate; ID50, half-maximal inhibitory dilution; IgG, immunoglobulin; MBI, multiepitope bead-based immunoassay; Nabs, neutralizing antibodies; OCR, ocrelizumab; S1P, sphingosine 1-phosphate receptor modulators.

Samples were available to measure functional neutralizing antibody (Nabs) titers in 77 patients with prior SARS CoV-2 infection. Nab levels showed a strong correlation with anti-RBD antibody levels detected by MBI assay (r=0.71, p<0.001), yet 21% of patients with high levels of binding antibodies did not have detectable functional Nabs. Compared to untreated patients, Nab titers were marginally lower in OCR-treated (p=0.055), and higher in Natalizumab-treated patients (p=0.01) (Figure 2C).

For each patient with symptomatic COVID-19 after OCR infusion, we plotted in Figure 3 the timeline of the last OCR infusion before infection (start of the gray bar); COVID-19 symptom onset (end of the grey bar); OCR infusion following infection if any (blue circle); and time of sampling (green rhomboid); alongside their respective Nab and anti-Spike MBI levels and TruCulture IFNγ responses. Of note, 23/27 (85%) patients who had an infection within six months after OCR infusion had low or undetectable (ID50 ≤100) Nab titers.

**Figure 3.**
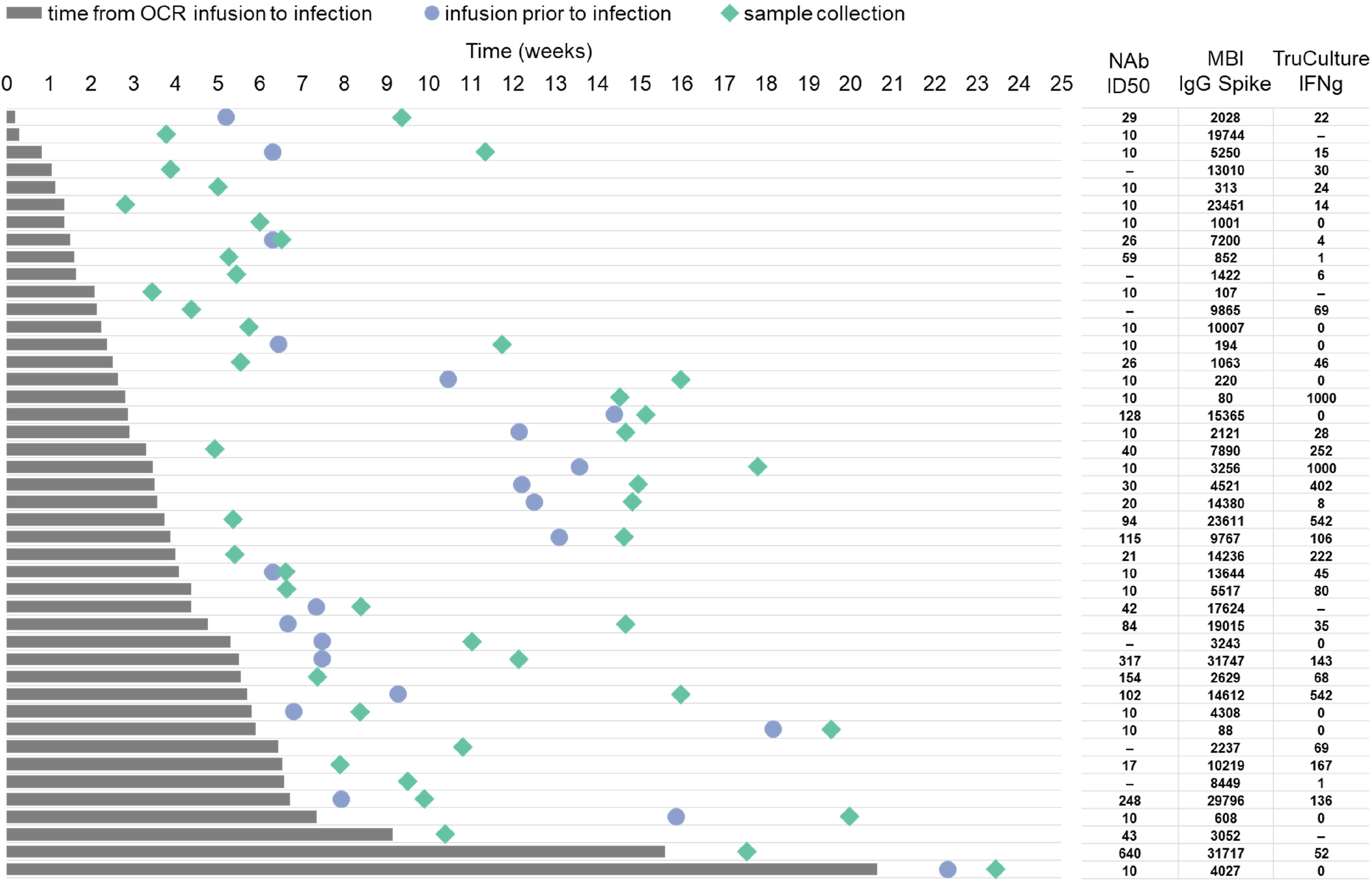
Timeline for symptomatic COVID-19 patients who were infected following OCR infusion. Legend: Timeline from last OCR infusion before infection (time zero, start of the grey bar) to COVID-19 infection onset (end of the grey bar), subsequent OCR infusion before sample collection (blue circle) and sample collection (green rhombus). Each line represents a patient timeline. Neutralizing Ab titers, binding IgG anti-spike level by MBI and TruCulture IFNγ are shown for each patient in the respective line. Neutralizing antibody titers are shown as log10 of half-maximal inhibitory dilution (ID50).

Multivariate analyses of all SARS-CoV-2 infected patients showed that treatment with OCR and the absence of hypertension predicted lower Elecsys antibody (the biological significance of the latter correlation is unknown). In symptomatic COVID-19 patients, treatment with OCR and longer infection-to-collection time correlated with lower MBI levels. In the subset of previously SARS-CoV-2 infected patients who were tested for Nabs, only OCR treatment at the time of infection was a predictor of lower Nabs titers, though the number of available samples for non-OCR DMTs was limited, e.g., only 5 samples for S1P.

### 6. Cellular responses to SARS-CoV-2 antigens in patients with prior SARS CoV-2 infection and in SARS-CoV-2 seronegative patients

We performed in-vitro T-cell stimulation studies on 159 of the 177 patients with prior SARS-CoV-2 infection (as defined in Section 3) using the TruCulture assay. Samples were not available or failed quality assurance check for 18 previously infected patients. Positive T-cell responses (above-zero values for both IFNγ and IL-2) were observed in 62/159 (39%) of all patients with prior infection. In the subset of patients for whom IFNγ responses were tested by both TruCulture and ELISpot tested (including one patient with missing TruCulture value), concordant positive rate was 59.6% (i.e. 53/89 samples tested positive for IFNγ on both TruCulture and ELISpot); concordant negative rate was 15.7% (14/89 samples tested negative on both assays); and discordant rate 24.7% (22/89 were positive for IFNγ on TruCulture and negative on ELISpot, or vice versa). In the subset with both TruCulture and ELISpot IL-2 responses tested, concordant positive rate 36.9% (31/84 were positive for IL-2 on both assays); concordant negative rate 33.3% (28/84); and discordant rate between TruCulture and ELISPot was 29.8% (25/84 samples). Cellular responses did not differ by race/ethnicity by either TruCultre or ELISpot (Supplemental Table).

We also performed in vitro T-cell stimulation studies on 130 MBI-seronegative patients who did not meet our criteria for infection. TruCulture reactivity was observed in 14/130 (11%) of the MBI-seronegative patients. None of these 14 patients were seropositive by Elecsys assay, nor by MBI Spike or RBD, but 9/14 (65%) were seropositive by NTD (non-receptor binding domain of Spike) by MBI and 2/4 (50%) had positive ELISpot results.

### 7. SARS-CoV-2 specific cellular responses in patients with prior COVID-19 infection stratified by DMT

OCR group had similar TruCulture IFNγ responses compared to the untreated reference group, while S1P showed depressed responses (p=0.0285) and Natalizumab had elevated responses (p=0.0002) (Figure 4A). IL-2 responses by TruCulture were also elevated with Natalizumab (p<0.0001), but were similar for all other DMTs, including OCR (Figure 4B). T-cell responses assessed by ELISpot are presented in Figure 4C for IFNγ and Figure 4D for IL-2. Only Natalizumab-treated patients had marginally elevated IL-2 responses (p=0.048), while S1P had marginally depressed IL-2 responses by ELISpot.

**Figure 4:**
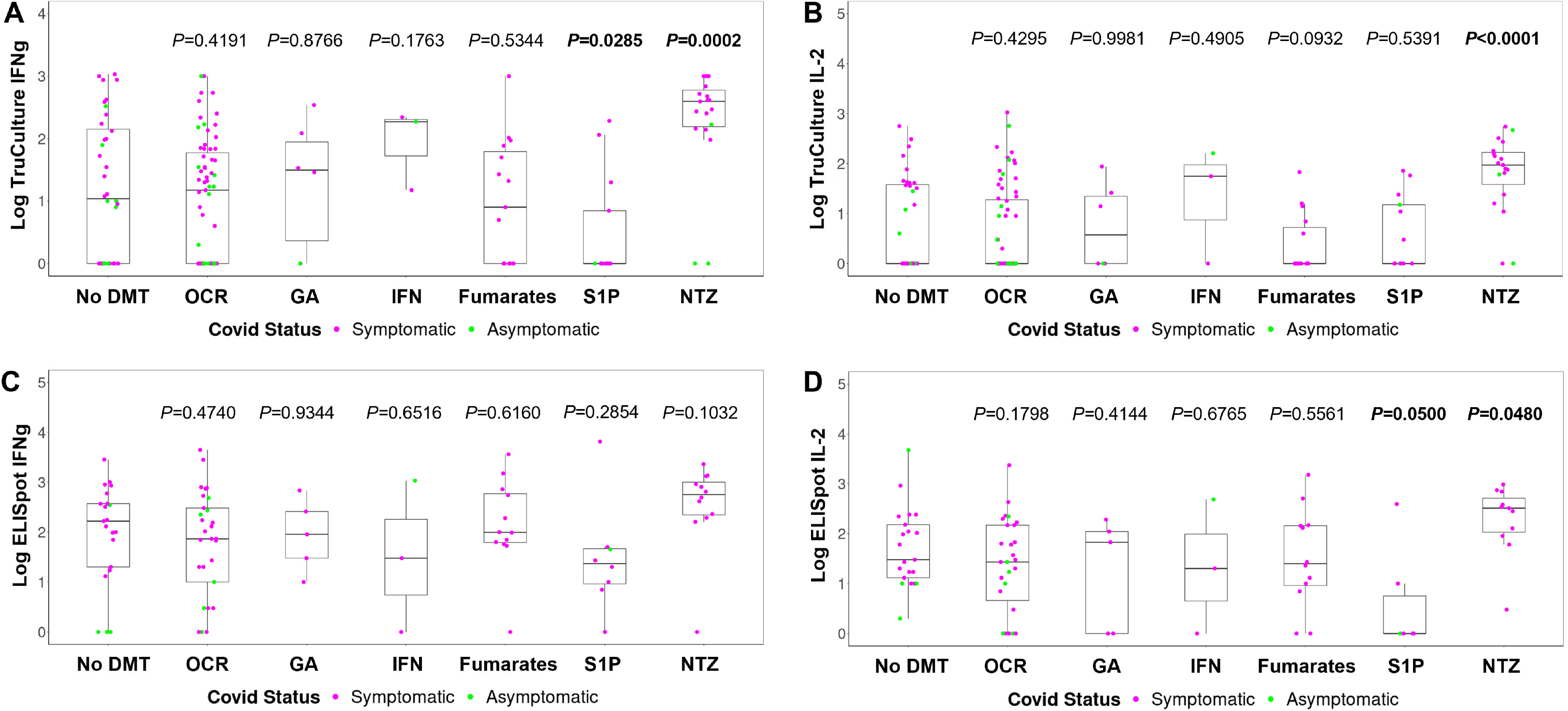
Induced T-cell activation in MS patients grouped by DMT based on IFNγ and IL2 secretion in TruCulture system (A and B) or evaluated by ELISpot (B and C). Legend: Symptomatic cases are shown in magenta and asymptomatic cases in green. P-values <0.05 are shown in bold.: Abbreviations: DMT, disease-modifying therapy; GA, glatiramer acetate; NAT, natalizumab; OCR, ocrelizumab; S1P, sphingosine 1-phosphate receptor modulators

In patients with prior SARS-CoV-2 infection, there was no trend for decreasing cellular responses (TruCulture IFNγ with increasing time from infection neither in the entire cohort nor in OCR subset (data not shown). The multivariate analyses did not identify any predictors of TruCulture responses.

In SARS-CoV-2 infected patients, anti-Spike antibody by MBI and cellular IFNγ responses by TruCulture showed a moderate degree of correlation overall (r=0.53, p<0.0001), and in both OCR (r=0.45, p=0.0002) (Figure 5A) and non-OCR (r=0.64, p<0.0001) (Figure 5B) subsets.

**Figure 5:**
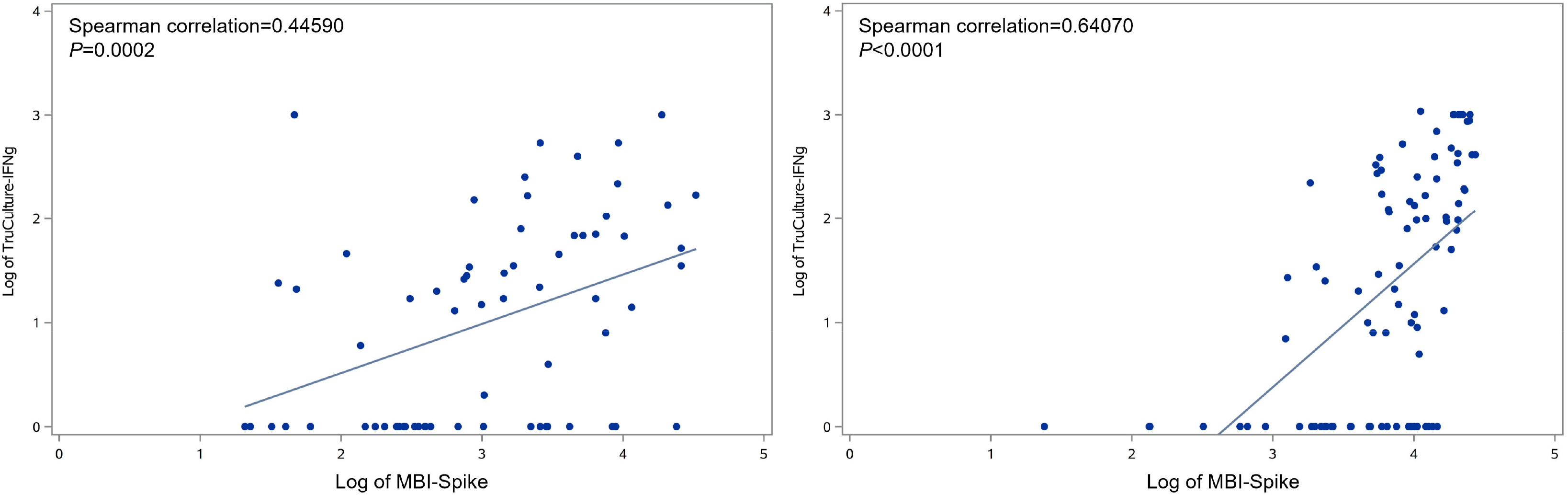
Correlation between TruCulture-IFNγ with MBI spike among patients with COVID receiving OCR (A) and receiving non OCR (B)

### 8. Relationship between COVID-19 infection symptoms and immune responses to SARS-CoV-2 in patients on OCR and other DMTs

In a multivariate model to predict MBI Spike levels based on DMT status and COVID-19 clinical variables (symptom duration, symptom number, and presence/absence of respiratory symptoms), only OCR treatment was a predictor for lower MBI Spike values. In a multivariate model to predict T-cell responses with TruCulture assay, longer COVID-19 symptom duration was associated with lower T-cell responses, but this relationship was driven by few outliers with ‘long COVID’ and was not present if patients with symptoms that persisted for >1 month were excluded. In the nine hospitalized patients, the mean anti-SARS-CoV-2 antibody values and T-cell responses were similar to the non-hospitalized group, except for TruCulture IFNγ responses that were higher in the hospitalized patients (data not shown).

## Discussion

In an ethnically-diverse group of 389 MS patients from the NYU MS Care Center in New York City, 46% had laboratory evidence of prior SARS-CoV-2 infection. This prevalence is higher than what would be expected for our area based on the NYC Department of Health seroprevalence study from July 2021 (the end of our study period),^29^ possibly due to over-representation in our Center of patients from Brooklyn, Queens, and Bronx neighborhoods with a very high incidence of prior SARS-CoV-2 infections (40-50%); use of highly sensitive multiplex bead-based immunoassays to measure seroprevalence; and the presumed greater motivation to participate in the study on the part of patients with suspected or known prior COVID-19. We confirmed COVID-19 diagnosis in 38% of patients with a history of COVID-19-like illness, but no commercial laboratory confirmation prior to enrollment, which is almost identical to the rate of SARS CoV-2 seropositivity among undocumented cases in a population-based study from New York City.^38^ The rate of asymptomatic infection in our patients - 27% - is lower than 33% rate in two large European studies, but higher than the 16% among World Trade Center responders in New York City area.^39^ Our results suggest that asymptomatic SARS CoV-2 infections are not uncommon among MS patients and occur at a rate comparable to the general population.

The high prevalence of SARS-CoV-2 infection in our MS patients allowed us to investigate how the immune memory to SARS-CoV-2 varies depending on DMT status at the time of infection. Patients who developed the infection while on OCR had an approximately 10-fold decrease in anti-SARS-CoV-2 binding antibody levels compared to the untreated group even though the time from infection to sample collection was, on average, 13 weeks shorter for OCR patients than all others. In multivariate analyses, OCR treatment was the single most important predictor of lower binding Ab levels. Functionally important neutralizing antibodies (Nabs) were also depressed with OCR compared to the untreated group, and 85% of patients who had an infection within six months of OCR infusion generated no or very low levels of Nabs.

On the other hand, anti-viral cellular responses, assessed via the TruCulture assay and by ELISpot (for a subset of patients) were present at a similar rate in OCR and untreated patients. T cell responses were largely independent of time from the last OCR infusion prior to infection, unlike humoral responses that tended to be weaker with shorter infusion-to-infection time. The uncoupling of antibody and T-cell responses in peripherally B-cell depleted patients, recently reported by others as well, ^20, 21^ may be due to the relative sparing of B-cells in secondary lymphoid organs, where T cell activation occurs, or to the fact that antigen-presenting function of memory B-cell is not essential for generating appropriate T-cell responses following COVID-19 infection or vaccination. Robust T-cell response has been associated with less severe disease^40–42^ and may in part explain ‘clinical-serologic dissociation’ in our mostly young and nondisabled patients: the similarity in COVID-19 severity across DMTs, despite markedly lower antibody responses with OCR compared to other DMTs.

Taken together, our data suggests that previously infected or vaccinated patients on aCD20 are less likely to be protected against ‘breakthrough’ infection than others, a prediction born out by a recent population-based study from UK,^43^ but are probably still protected against COVID progression due to intact T cell immunity.^15, 18, 44, 45^ Memory T cells that contribute to protection against severe disease by eliminating infected cells and limiting viral replication may explain, at least in part, why vaccines prevent hospitalizations and death even against variants that exhibit limited neutralization by vaccine-induced humoral immunity.^46, 47^ Indeed, T cells responses to Beta and Omicron variants have been documented following the third dose of mRNA vaccines even in aCD20-treated patients.^48^

Patients who experienced SARS-CoV-2 infection while on S1P receptor modulators tended to have depressed humoral and, to a larger extent, cellular immune responses than untreated patients. This may be a consequence of peripheral lymphopenia, disruption of immune cell traffic to and from the germinal center, or the impact of the drug on the downstream signaling pathways involved in cytokine production. T-cell responses in S1P-treated patients are also reduced following COVID-19 vaccination.^1, 44, 45^ In another example of clinical-immunologic dissociation, COVID-19 outcomes were not worse in patients on S1P in our group or other large series.^9, 49^ A possible explanation is that S1P modulators may help protect against immune system hyperactivation — ‘cytokine storm’ - and stabilize pulmonary epithelium,^50^ thereby preventing more severe disease.

Natalizumab-treated patients antibody and cellular responses were on par, or better, than in untreated patients, possibly as a consequence of elevated T- and B-cell lymphocyte counts in the peripheral circulation with this therapy.^51^ In patients treated with fumarates, both humoral or cellular responses were intact despite the lower peripheral lymphocyte counts with these drugs.^52^ In contrast to a prior study,^20^ we did not observe impairment in immune responses in IFNβ-treated patients, in line with the observation from large cohorts that COVID-19 outcomes are actually better in IFNβ –treated patients than with other DMTs.^9^ In patients on glatiramer and teriflunomide, immune responses were intact though firm conclusions cannot be made given the relatively small number of infections in each of these DMT classes (<11 patients per DMT groups).

Several strengths of our study deserve mention. First, the large number of studied patients and high SARS-CoV-2 seropositive rate allowed for statistically meaningful comparisons of immunologic outcomes for the different DMTs.^20^ Secondly, our population largely reflects the diversity of New York City, with 29% of patients self-identifying as African-Americans and 27% - as Hispanics. (Asians were relatively underrepresented in our Center, likely due to the lower prevalence of MS in this ethnic group.) In univariate and multivariate analyses, race/ethnicity was not a predictor of clinical outcomes, nor of antibody or cellular anti-SARS-CoV-2 responses in either OCR or non-OCR patients (Supplemental Table 1). Third, due to the epidemiology of COVID-19 spread in our area — with a highly destructive first wave in Spring of 2020 followed by a large second wave in Fall-Winter 2020 — we were able to collect samples with a wide range of time-to-infection and investigate the durability of immune responses over the median interval of 43 week from infection (interquartile range: 14-50 week). Fourth, we used a custom Multiepitope Bead-based Immunoassay (MBI) specially designed to interrogate immune responses to SARS-CoV-2 antigens with enhanced sensitivity. MBI was instrumental in identifying serologic evidence of infection in OCR-treated patients with depressed antibody responses and asymptomatic infections, which were often missed with the commercially available Elecsys test designed for high throughput in the clinical setting. Differences in seropositivity rates in OCR patients with past infection (36% by Elecsys and 89% by MBI) emphasize the importance of considering assay sensitivity and the clinical context when interpreting published seroprevalence studies. The higher sensitivity of multiplex bead array over electro-chemiluminescence immunoassay has been noted by others as well.^20^ Fifth, we assessed functional neutralizing responses with live virus microneutralization assays in a subset of patients with binding antibodies to SARS-CoV-2. Despite a high degree of correlation between binding (MBI IgG Spike) and neutralizing antibody levels (r=0.71), one in five patients with binding antibodies had low or undetectable neutralizing antibodies. This finding calls into question over-reliance on commercially available antibody binding assays as surrogates of serologic immunoprotection. Sixth, we used a technically simple, rapid test of cellular responses to SARS-CoV-2 – TruCulture assay – and showed its utility even in patients with suppressed humoral immunity. Because TruCulture is a relatively new assay that has only been used in a few published studies,^53^ we corroborated TruCulture results with a conventional but more technically demanding ELISpot in a subset of patients. The two tests were largely concordant, though slightly better for for IFNγ responses – less than 25% of patients had discordant results by the two assays. Specificity of TruCulture merits further investigation. We identified a group of 14 out of 130 (11%) patients who were positive on TruCulture, but serologically negative by MBI. These TruCulture-positive, seronegative patients may represent false positives (possibly due to cross-reaction with other coronaviruses), or true-positives, with absent antibody titers during convalescence,^42^ or ‘aborted infection’ in which antibody responses fail to develop in the first place.^54^

Limitations of our study include the lack of SARS-CoV-2 PCR confirmation for all of our patients, especially those infected during the first wave, in which 80% of patients were not PCR-confirmed.^55^ Our inclusion criteria, though intentionally broad, precluded older, systemically immunocompromised, severely disabled patients from participation. Patients who were lost due to fatal COVID-19 infection could not be accounted for in our study. Our patients largely reflect the mild range within the spectrum of SARS-CoV-2, as evidenced by the fact that only 7% of our symptomatic cases were hospitalized as compared to the average hospitalization rate of 16% across MS studies ^56^. Our findings may not be generalizable to older and more disabled patients, who account for the bulk of excess morbidity and mortality in MS.^57^

The main conclusion of our study is that relatively young and otherwise healthy MS patients generally had favorable clinical course across DMTs despite markedly impaired adaptive immune responses associated with some of the DMTs (aCD20, S1P). To better understand the uncoupling of T-cell from antibody responses in aCD20 treated patients and to identify predictors of immune response in patients on the different DMTs, we are conducting in-depth immunophenotyping and activation-induced marker studies. Race/ethnicity did not predict either clinical or immunologic outcomes.

## Data Availability

All data produced in the present study are available upon reasonable request to the authors and review and approval by the study sponsor and collaborator.

## Acknowledgments

The authors wish to gratefully acknowledge our medical assistants, Ms. Apphia Daniel, Ms. Teodora Drakulovic, Ms. Rosetta Ellis, for their gentle bedside manner and great phlebotomy skills. We are grateful to all of our colleagues who assisted us with patient recruitment and provided feedback on our study and to all the patients who participated in this study.

---

**Supplemental Figure 1:** MBI Spike, RBD and NTD in healthy, pre-pandemic controls (’Healthy’) and confirmed, non-MS COVID-19 cases (’COVID+’)

Abbreviations: MBI, multiepitope bead-based immunoassay; NTD, N-terminal domain; RBD, receptor-binding domain; S, Spike. MFI, mean fluorescence intensity (MFI)

**Supplemental Figure 2:** Distribution of COVID cases by calendar time

Legend: Distribution of COVID-19 cases by calendar months reflects the biphasic distribution of cases in our area (https://covidactnow.org/us/metro/new-york-city-newark-jersey-city_ny-nj-pa/?s=25581470). First COVID-19 cases were reported in our area in February 2020. No patients in our group had COVID between May 2021 and July 2021 (the last collection date).

## Notes

Acknowledgments: This work was supported by an unrestricted investigator-initiated grant from Genentech. Editorial assistance for the figures and tables, furnished by Sarah Nordquist, PhD, of Health Interactions, Inc, was provided by F. Hoffmann-La Roche Ltd.

### Competing Interest Statement

IK served on the scientific advisory board for Biogen Idec, Genentech, Alexion, EMDSerono; received consulting fees from Roche; and received research support from Guthy-Jackson Charitable Foundation, National Multiple Sclerosis Society, Biogen Idec, Serono, Genzyme, and Genentech/Roche; he receives royalties from Wolters Kluwer for 'Top 100 Diagnosis in Neurology' (co-written with Jose Biller)
GJS received honoraria from BMS, Eli Lilly and Genentech, and research support from BMS, Genentech, Lupus Research Alliance, NIH-NIAMS, NIH-NIAID and NIH-NILB.
MK is on the scientific advisory board for NexImmune and Genentech and received research support from Merck Sharp & Dohme Corp., a subsidiary of Merck & Co., Inc., Genentech, the Mark Foundation, NIH-NIGMS and NIH-NCI.
CR and RP are employees and shareholders of F. Hoffmann-La Roche.
MJM reported the following potential competing interests: laboratory research and shareholder of F. Hoffmann-La Roche Ltdclinical trials contracts for vaccines or MAB vs SARS-CoV-2 with Lilly, Pfizer, and Sanofi; personal fees for Scientific Advisory Board service from Merck, Meissa Vaccines, and Pfizer; contract funding from USG/HHS/BARDA for research specimen characterization and repository; research grant funding from USG/HHS/NIH for SARS-CoV-2 vaccine and MAB clinical trials.
LZR served on the scientific advisory board for Biogen, Genentech, Celgene, and Novartis and received research support from Consortium of Multiple Sclerosis Centers; Biogen; and Genentech.
MC is an employee and shareholder of Genentech, Inc.
KH is a former employee of Genentech, Inc.
JP is an employee of Genentech, Inc. and shareholder of F. Hoffmann-La Roche.
TEB, RC, ZR, KP, SE, YY, AS have nothing to disclose.

### Funding Statement

This work was supported by an unrestricted investigator-initiated grant from Genentech. Editorial assistance for the figures and tables, furnished by Sarah Nordquist, PhD, of Health Interactions, Inc, was provided by F. Hoffmann-La Roche Ltd.

### Author Declarations

The Institutional Review Board (IRB) of NYU Langone Health gave ethical approval for this work.

