## Supplemental Table for "Cellular and humoral immunity to SARS-CoV-2 infection in multiple sclerosis patients on ocrelizumab and other disease-modifying therapies: a multi-ethnic observational study"

**Supplementary Table.** Clinical characteristics of MS patients with lab-confirmed COVID receiving OCR vs Non-OCR by Race/ Ethnicity

|  | **OCR** | | | | | **Non-OCR** | | | | |
| --- | --- | --- | --- | --- | --- | --- | --- | --- | --- | --- |
|  | **White n=27** | **AA n=14** | **HA n=23** | **Other n=6** | **Total**  **n=70** | **White n=42** | **AA n=35** | **HA n=26** | **Other n=5** | **Total**  **n=107** |
| Age, years |  |  |  |  |  |  |  |  |  |  |
| Mean (SD) | 42.2 (9.6) | 37.6 (8.7) | 34.4 (7.4) | 32.3 (13.3) | 37.9  (9.7) | 43.9 (11.4) | 41.1 (10.9) | 37.8 (10.8) | 37.3 (17.2) | 41.3  (11.4) |
| Sex, n (%) |  |  |  |  |  |  |  |  |  |  |
| Female | 22 (81.5) | 9 (64.3) | 17 (73.9) | 5 (83.3) | 53  (75.7) | 20 (47.6) | 29 (82.9) | 19 (73.1) | 4 (100) | 72  (67.3) |
| Ambulatory status, n (%) |  |  |  |  |  |  |  |  |  |  |
| Fully ambulatory | 23 (85.2) | 8 (57.1) | 14 (60.9) | 5 (83.3) | 50  (71.4) | 31 (73.8) | 22 (62.9) | 22 (84.6) | 2 (50.0) | 77  (72.0) |
| COVID Symptom count, n (%) |  |  |  |  |  |  |  |  |  |  |
| 0 | 5 (18.5) | 3 (21.4) | 8 (34.8) | 3 (50.0) | 19 (27.1) | 7 (16.7) | 11 (31.4) | 5 (19.2) | 1 (25.0) | 24 (22.4) |
| 1-3 | 5 (18.5) | 7 (50.0) | 4 (17.4) | 1 (16.7) | 17 (24.3) | 8 (19.0) | 4 (11.4) | 10 (38.5) | 2 (50.0) | 24 (22.4) |
| >3 | 17 (63.0) | 4 (28.6) | 11 (47.8) | 2 (33.3) | 34 (48.6) | 27 (64.3) | 20 (57.2) | 11 (42.3) | 1 (25.0) | 59 (55.2) |
| Respiratory symptoms, n (%) | 5 (18.5) | 3 (21.4) | 8 (34.8) | 0 | 16 (22.9) | 16 (38.1) | 11 (31.4) | 11 (42.3) | 0 | 38 (35.5) |
| Hospitalization, n (%) | 1 (3.7) | 1 (7.1) | 0 | 0 | 2 (2.9) | 4 (9.5) | 2 (5.7) | 1 (3.8) | 0 | 7 (6.5) |
| Infection to collection, weeks |  |  |  |  |  |  |  |  |  |  |
| n | 22 | 7 | 12 | 3 | 44 | 31 | 23 | 15 | 3 | 72 |
| Mean (SD) | 26.8 (19.0) | 23.2 (16.3) | 26.2 (19.0) | 36.0 (22.4) | 26.7  (18.4) | 36.6 (18.4) | 45.6 (13.5) | 41.3 (17.1) | 6.9 (1.0) | 39.2 (17.9) |
| Median (Q1, Q3) | 16.9 (9.6, 46.1) | 13.7 (12.0, 42.4) | 20.1 (10.9, 46.1) | 43.0 (11.0, 54.1) | 18.0 (11.1, 45.6) | 44.7 (15.0, 51.4) | 49.1 (44.1, 53.0) | 47.9 (24.7, 51.6) | 6.4 (6.1, 8.0) | 46.4 (19.8, 51.6) |
| Log of Elecsys anti-SARS-CoV-2 Ab |  |  |  |  |  |  |  |  |  |  |
| n | 26 | 14 | 23 | 6 | 69 | 36 | 29 | 23 | 4 | 92 |
| Mean (SD) | 0.3^a^ (0.6) | 0.5 (0.9) | 0.3 (0.5) | 0.0 (0.0) | 0.3 (0.6) | 1.6^b^ (0.9) | 1.9^c^ (0.8) | 1.4^d^ (1.1) | 1.5 (1.1) | 1.6 (0.9) |
| Median (Q1, Q3) | 0.0 (0.0, 0.8) | 0.0 (0.0, 0.6) | 0.0 (0.0, 0.6) | 0.0 (0.0, 0.0) | 0.0  (0.0, 0.6) | 1.7 (0.8, 2.2) | 2.1 (1.4, 2.3) | 1.6 (0.0, 2.2) | 1.7 (0.5, 2.4) | 1.8  (1.0, 2.2) |
| Log of MBI IgG Sspike Ab |  |  |  |  |  |  |  |  |  |  |
| n | 27 | 14 | 23 | 6 | 70 | 42 | 35 | 26 | 4 | 107 |
| Mean (SD) | 3.0 (0.9) | 3.3 (0.9) | 3.0 (0.9) | 2.7 (0.3) | 3.0  (0.9) | 3.7 (0.5) | 3.9 (0.4) | 3.6 (0.7) | 3.5 (1.4) | 3.7  (0.6) |
| Median (Q1, Q3) | 3.0 (2.5, 3.7) | 3.4 (2.4, 4.3) | 3.2 (2.4, 3.9) | 2.6 (2.5, 3.0) | 2.6  (2.5, 3.0) | 3.8 (3.4, 4.1) | 4.0 (3.7, 4.3) | 3.8 (3.3, 4.0) | 4.1 (2.6, 4.3) | 3.9  (3.4, 4.2) |
| Log of Nabs ID50 |  |  |  |  |  |  |  |  |  |  |
| n | 16 | 5 | 9 | 2 | 32 | 22 | 11 | 10 | 2 | 45 |
| Mean (SD) | 1.3^e^ (0.5) | 1.5^f^ (0.7) | 1.6^g^ (0.5) | 1.0^h^ (0.0) | 1.4  (0.5) | 2.1^i^ (0.7) | 1.8^j^ (0.8) | 1.8^k^ (0.3) | 2.5^l^ (0.1) | 2.0  (0.6) |
| Median (Q1, Q3) | 1.0 (1.0, 1.5) | 1.0 (1.0, 1.9) | 1.4 (1.3, 2.0) | 1.0  (1.0, 1.0) | 1.1  (1.0, 1.8) | 2.4 (1.6, 2.5) | 1.9 (1.4, 2.3) | 1.9 (1.6, 2.0) | 2.5 (2.4, 2.5) | 1.9  (1.6, 2.5) |
| Log of TruCulture IFNγ |  |  |  |  |  |  |  |  |  |  |
| n | 25 | 14 | 22 | 6 | 67 | 35 | 29 | 24 | 4 | 92 |
| Mean (SD) | 1.2 (0.9) | 0.8 (1.2) | 1.2 (1.0) | 0.3 (0.5) | 1.0 (1.0) | 1.4 (1.1) | 1.5 (1.1) | 1.0 (1.2) | 1.4 (1.6) | 1.3 (1.1) |
| Median (Q1, Q3) | 1.3  (0.0, 1.7) | 0.0  (0.0, 1.9) | 1.3  (0.0, 1.8) | 0.0  (0.0, 0.3) | 1.2  (0.0, 1.8) | 1.7  (0.0, 2.4) | 1.5  (0.8, 2.3) | 0.0  (0.0, 1.9) | 1.3  (0.0, 2.8) | 1.4  (0.0, 2.4) |
| Log of TruCulture IL2 |  |  |  |  |  |  |  |  |  |  |
| n | 25 | 14 | 22 | 6 | 67 | 35 | 29 | 24 | 4 | 92 |
| Mean (SD) | 0.5 (0.8) | 0.8 (1.1) | 0.7 (0.9) | 0.3 (0.6) | 0.6 (0.9) | 0.9 (1.0) | 1.0 (1.0) | 0.8 (0.9) | 0.8 (0.8) | 0.9 (0.9) |
| Median (Q1, Q3) | 0.0  (0.0, 1.0) | 0.0  (0.0, 1.7) | 0.2  (0.0, 1.5) | 0.0  (0.0, 0.0) | 0.0  (0.0, 1.3) | 1.0  (0.0, 1.7) | 1.2  (0.0, 1.9) | 0.0  (0.0, 1.7) | 0.5  (0.2, 1.3) | 0.7  (0.0, 1.8) |

Ab, antibodies; AA, African American; HA, Hispanic American; OCR, ocrelizumab.
