## Supplementary figures and images for "Cellular and humoral immunity to SARS-CoV-2 infection in multiple sclerosis patients on ocrelizumab and other disease-modifying therapies: a multi-ethnic observational study"

### Supplemental Figures

**Supplemental Figure 1.**

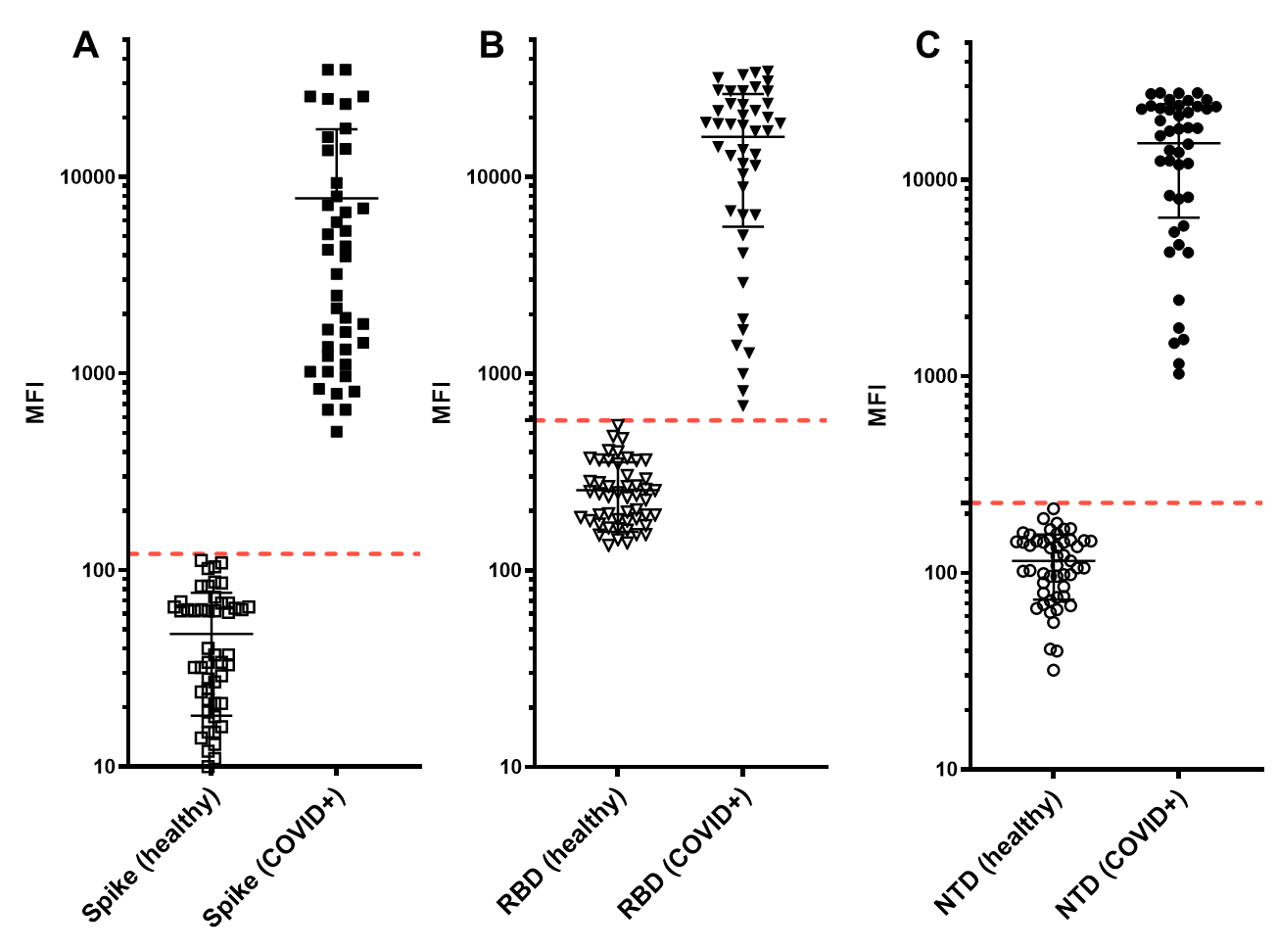

**Supplemental Figure 2.**

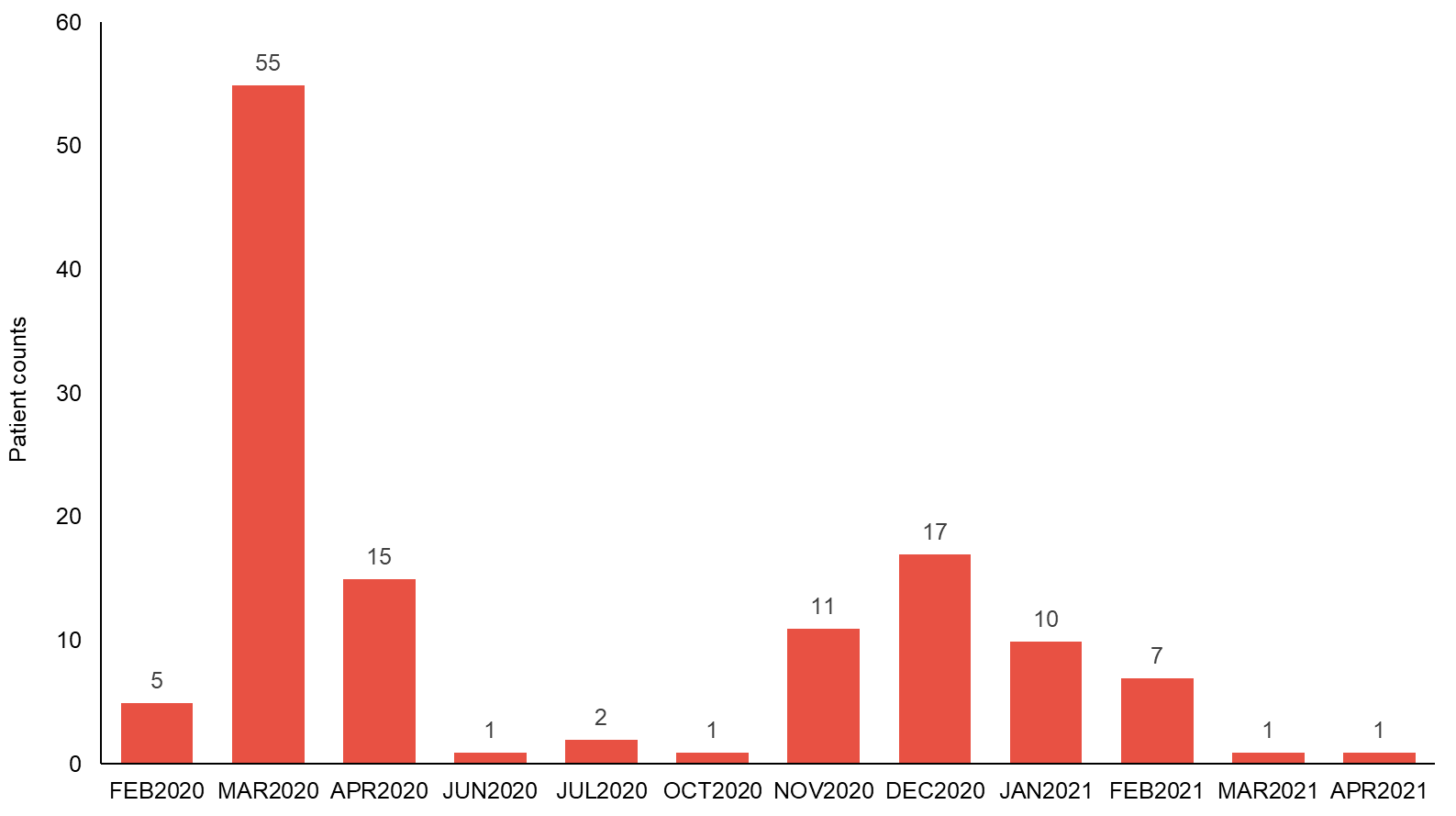
